# Discovering Social Determinants of Health from Case Reports using Natural Language Processing: Algorithmic Development and Validation

**DOI:** 10.1101/2022.11.30.22282946

**Authors:** Shaina Raza, Elham Dolatabadi, Nancy Ondrusek, Laura Rosella, Brian Schwartz

## Abstract

**Background:** Social determinants of health are non-medical factors that influence health outcomes (SDOH). There is a wealth of SDOH information available in electronic health records, clinical reports, and social media data, usually in free text format. Extracting key information from free text poses a significant challenge and necessitates the use of natural language processing (NLP) techniques to extract key information.

**Objective:** The objective of this research is to advance the automatic extraction of SDOH from clinical texts.

**Setting and Data:** The case reports of COVID-19 patients from the published literature are curated to create a corpus. A portion of the data is annotated by experts to create ground truth labels, and semi-supervised learning method is used for corpus re-annotation.

**Methods:** An NLP framework is developed and tested to extract SDOH from the free texts. A two-way evaluation method is used to assess the quantity and quality of the methods.

**Results:** The proposed NER implementation achieves an accuracy (F1-score) of 92.98% on our test set and generalizes well on benchmark data. A careful analysis of case examples demonstrates the superiority of the proposed approach in correctly classifying the named entities.

**Conclusions:** NLP can be used to extract key information, such as SDOH factors from free texts. A more accurate understanding of SDOH is needed to further improve healthcare outcomes.

## INTRODUCTION

### Background and significance

Social determinants of health (SDOH) refer to the non-medical factors such as birth, education, occupation, and living conditions, that influence the health outcomes of individuals [1]. There is extensive evidence [2–5] that SDOH significantly influences a broad range of health outcomes including mortality rate, elderly care, mental health, and risks for chronic diseases such as asthma, cancer, heart disease, and obesity [6]. Diabetes, depression, hypertension, and suicidal behavior are all outcomes of SDOH [7]. According to some studies, medical care accounts for 10-20% of health factors, while SDOH accounts for 80% to 90% [8]. Thus, it is of high importance to address the SDOH to improve the health systems.

SARS-CoV-2 is a virus that infects humans, causing severe upper respiratory problems and coronavirus disease 2019 (COVID-19) disease [9]. Recent statistics suggest that SDOH, such as race, ethnicity, gender, social-economic factors, and related population characteristics are also among the risk factors for COVID-19 [10]. There is evidence [11, 12] that those who are homeless have a higher prevalence of COVID-19 disease than those who are housed. As a result, understanding the SDOH in the context of a pandemic is critical for improving population health outcomes.

Despite advancements in technology, the collection of SDOH data remains a challenge for the public health community. Electronic Health Records (EHRs) constitute a significant portion of clinical data [13], but their use in clinical research is limited due to difficulties in automating the process of extracting information from unstructured data. Natural Language Processing (NLP) techniques can potentially overcome these limitations by facilitating the extraction of relevant information from unstructured data, thereby enabling its use in analysis and decision-making.

Through this work, we try to address the research question “How can we effectively mine SDOH data from case reports to improve its practical usability and scholarly value?”

### Objectives

The objectives of this study are:

- To propose an SDOH NLP framework that can extract SDOH information from case report data. This framework aims to provide a more comprehensive understanding of the patient’s condition by considering the impact of social and economic factors.
- To prepare a dataset that is annotated with SDOH labels. This dataset will be used for training and evaluating the NLP framework. In addition, semi-supervised learning techniques will be implemented to facilitate data re-annotation and improve the accuracy of the SDOH annotations in the dataset.

This work adds to previous efforts [14–17] by acknowledging both clinical and SDOH factors as key contributors to understanding patient health outcomes. The proposed framework includes a data processing module that parses the contents of free text and prepares a dataset, an NLP module that extracts SDOH information from the texts, and an evaluation module that assesses the NLP module’s accuracy and effectiveness.

Experimental results show that the proposed approach outperforms the baseline methods across multiple datasets. A thorough analysis of the extracted information yields important findings that can be utilized for COVID-19 surveillance and the creation of informed public health strategies.

## MATERIALS AND METHODS

As part of a research project, we developed an NLP framework and validated the performance of its module for SDOH identification. The proposed framework is extensible and applicable to a variety of public health use cases, including surveillance, epidemiology, and policy-making processes.

### Data

Our dataset is an extension and refinement of our original collection [18]. This enhanced version specifically highlights the SDOHs. We derived this data from 4,000 electronic case reports of COVID-19 patients obtained via the LitCOVID API [19], marking a shift in focus from purely clinical elements (as in previous version) to the broader context of SDOH. Our aim is to illuminate health disparities and the elements that drive them.

We thoroughly applied selection criteria to ensure data relevance and quality, such as a specific timeframe (January to June 2022), language (English), defined patient age groups, and the exclusive inclusion of peer-reviewed case reports. The search query is given in Appendix A: Table S1.

The SDOH has been conceptually organized into categories for a more holistic understanding of health outcomes among COVID-19 patients. These categories are given below, and more fine-grained details of named entities are in Appendix A: Table S2.

1. *Demographic Factors*: Gender, Age, Race/Ethnicity -These factors allow us to identify patterns and trends among different demographic groups.
2. *Biometric Factors:* Height, Weight -These physical characteristics impact an individual’s health status.
3. *Temporal Factors*: Date, Relative Date, Duration, Time -These provide insights into temporal trends such as disease progression and recovery time.
4. *Lifestyle Factors:* Smoking, Alcohol, Substance use -These lifestyle attributes can significantly impact health outcomes.
5. *Socioeconomic Factors: Employment* -This factor influences access to healthcare, stress levels, and other health-related factors.
6. *Healthcare System Interaction*: Admission/Discharge -These data points are essential for understanding the patient’s journey within the healthcare system and the efficiency of their care.
7. *Clinical Factors:* As detailed in Appendix A, Table S2 -These clinically relevant data points derived from patient records influence health outcomes. This dataset and categorization scheme offers a broader and more detailed understanding of the factors that influence the health outcomes of COVID-19 patients, with a specific emphasis on the SDOH.

### Proposed SDOH NLP Framework

The SDOH NLP framework, shown in Figure 1, features three modules: (1) data processing, (2) NLP, and (3) evaluation.

**Figure 1:**
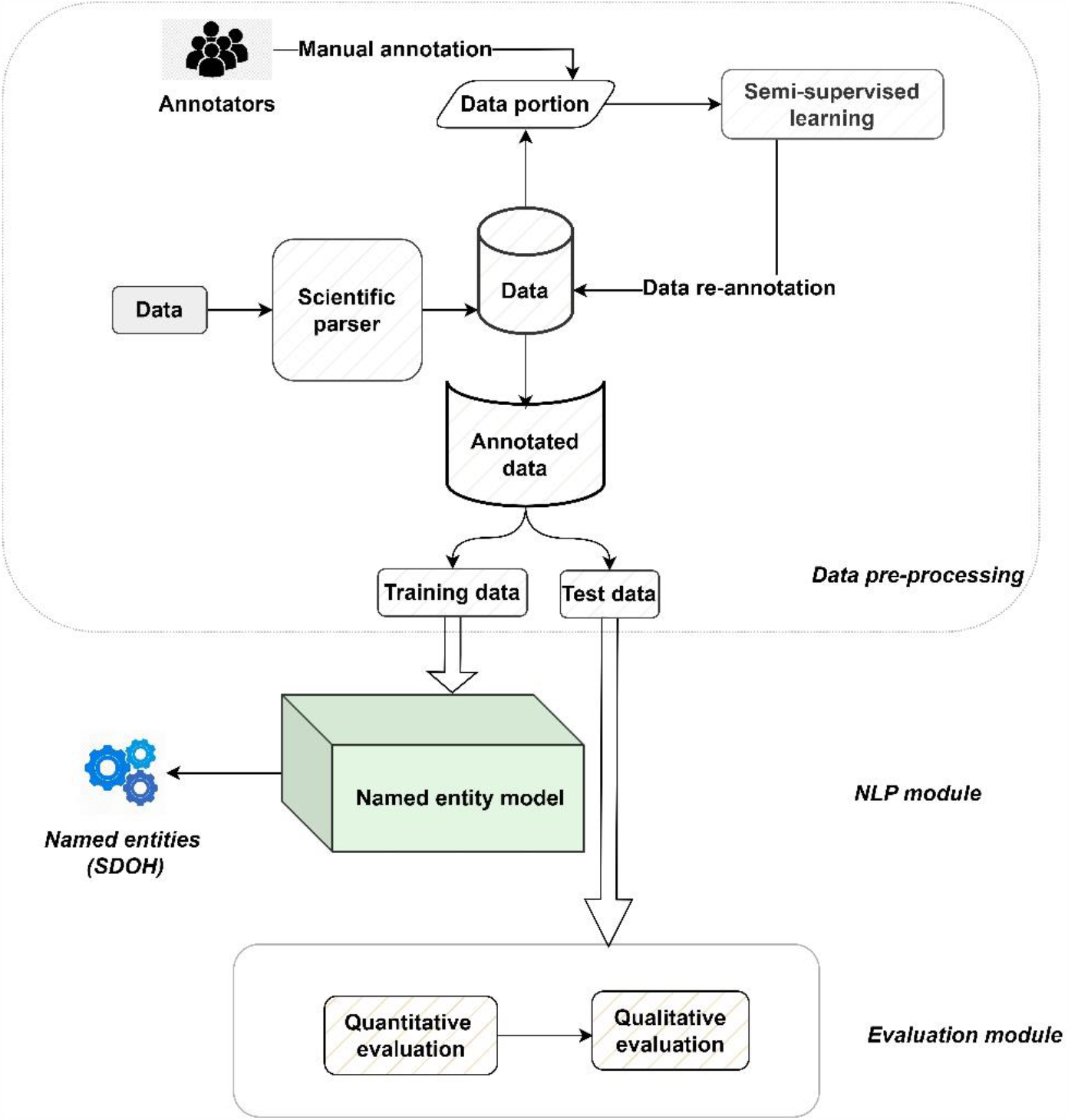
SDOH NLP proposed architecture

### Data processing module

Case reports, acquired via the LitCOVID API, were processed by a scientific parser software [20] to extract textual content and were stored in MongoDB database. Each database entry represents a case report, organized by a unique identifier. A group of four experts manually annotated a sample of 200 case reports, establishing the basis of our annotated dataset. This process is facilitated by the Spark NLP annotation tool [21] resulted in approximately 5000 sentences being annotated in three months, with each sentence has multiple SDOH categories in many instances.

To ensure annotation consistency, we adhered to literature-sourced guidelines [22, 23] for named entity annotation. The Inter-annotator Agreement (IAA) [24] approach was utilized to measure annotation alignment across all annotators, with the Cohen’s Kappa [25] coefficient providing a measure of agreement. We obtain the Cohen’s Kappa value of 0.75, which is classified as substantial agreement on the kappa coefficient scale [26]. Further, the discrepancies in annotation were resolved through a consensus-based dialogue between annotators.

Pre-processing of our data included standardization measures such as lowercasing all text, removing foreign or uncommon symbols, and separating contractions into individual words. We utilized tokenization to break the text down into manageable units and removed stop words to spotlight the salient terms. This refining process prepared our data for an efficient and in-depth analysis.

For large-scale annotation, we implemented a semi-supervised approach. Initial manual annotations (on 200 case reports) were used to train a BERT (Bidirectional Encoder Representations from Transformers) model for the NER task, achieving an accuracy of approximately 94.18%. In NER tasks, the model predicts the label of each token—usually a word—in a sentence. These labels represent categories or “entities,” such as person names, locations, medical codes, time expressions, and particularly, SDOH in our case. Next, we used this trained model to annotate the larger dataset (remaining 3800 reports), effectively using the model to classify or predict the NER label for each token in these reports. This process is also known as pseudo-labeling. The newly annotated data is then combined with our original labeled data, and the model is retrained on this combined dataset. This cycle can be repeated as necessary. The key idea is that the performance of the model is continuously improved as it learns from both the manually annotated data and the new data that it annotates itself.

Through our empirical analysis, we discovered that the dataset containing 4000 case reports encompasses approximately 60,000 sentences. Within this dataset, we further identified around 80,000 named entities related to SDOH and approximately 180,000 clinical named entities. The annotated data is stored in the widely used CONLL-2003 format [27], for NER tasks. The NER model, discussed in the NLP layer, is trained using this annotated data, with a portion (30%) reserved for evaluation purposes. While the foundational method is inspired by our earlier work [18], the training process for SDOH has undergone significant re-training in our current study.

The schema of this dataset version, we introduce is:

1. Sentence ID <INTEGER>: A unique identifier allocated to each sentence, maintained from the original version.
2. Word <STRING>: This represents the individual word token from the sentence, preserved from the original version.
3. POS <STRING>: The Part of Speech tag assigned to the word, also carried over from the original version.
4. Chunk <STRING>: The syntactic chunk tag of the word, kept consistent with the original version.
5. NER Tag <ENUM>: This version introduces an enhancement to the NER tag associated with each word. In addition to ‘Clinical’, this has been expanded to encompass ‘Non-Clinical’ and ‘SDOH’ categories. For multi-label classification, each word in a sentence would have a distinct named entity tag, allowing multiple labels per sentence.

For multi-label classification, each word in a sentence would have a distinct named entity tag, allowing multiple labels per sentence.

### Natural language processing (NLP) module

Our proposed framework utilizes an NLP method for Named Entity Recognition (NER) as a sequence classification task. Every token in the input sequence is assigned a label, for a more effective entity identification. An illustration of this would be assigning “O, I-PERSON, O, I-DISEASE” to “The patient has COVID-19”, where “O” represents a non-entity type and “I-DISEASE” a single-token DISEASE type.

The NER model integrates three key players: a Transformer layer, a Bidirectional Long Short-Term Memory (BiLSTM), and a Conditional Random Field (CRF) layer, as illustrated in Figure 2. Among these, the Transformer layer utilizes BioBERT [28], which has been specifically fine-tuned on our dataset for NER task. This task-specific adaptation transforms input sequences into detailed embeddings, providing robust representations tailored to our objectives.

**Figure 2:**
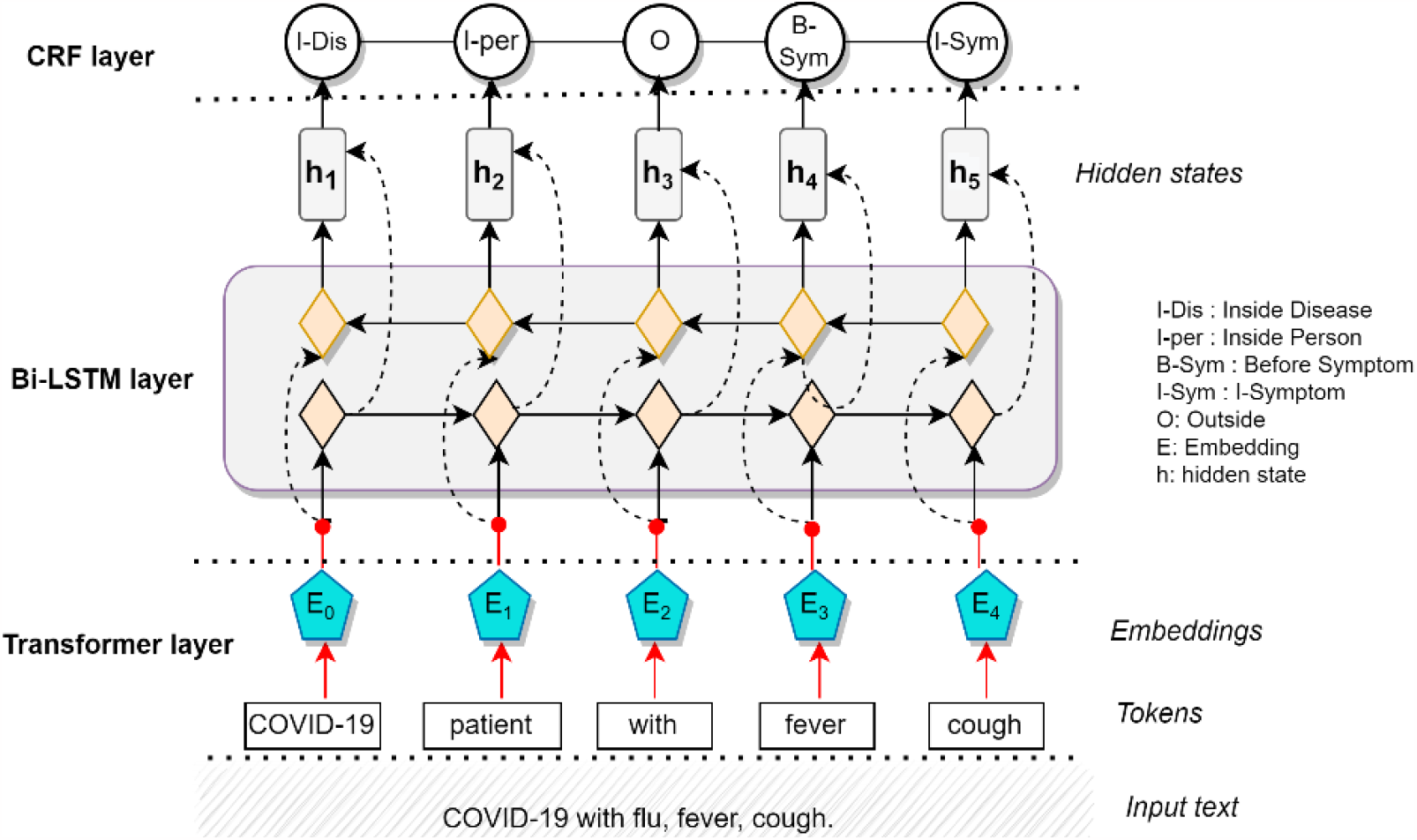
Proposed framework for named entities task.

Subsequent to the Transformer layer, the BiLSTM layer captures context features to amplify the semantic meanings in the texts. The layer’s forward and backward LSTM successfully bring together hidden information from both preceding and subsequent texts.

Finally, the CRF layer [29] inputs the output sequence from the BiLSTM layer, revealing the dependencies within it. This layer effectively translates the complex interdependency between the named tags into the final predicted labels. The final IOB representation, output from the CRF layer, is converted into an accessible format by linking the recognized named entities with their appropriate labels.

#### Joint optimization

In our NLP strategy, we initially fine-tune BioBERT for the task at hand using training data, which enables BioBERT to learn task-related patterns. Next, we combine BioBERT with the BiLSTM and CRF layers in a joint optimization approach. This means all layers, including BioBERT and subsequent layers (BiLSTM and CRF), are trained collectively, facilitating the optimization of the entire model architecture. By initially fine-tuning BioBERT and then jointly training it with BiLSTM and CRF layers, we exploit both BioBERT’s pre-existing knowledge and the task-oriented data captured by the following layers.

As an example, a few named entities annotated on a sample case report is shown in Appendix A: Figure S1.

### Evaluation module

In this study, a thorough evaluation protocol was adopted to assess the efficacy of our proposed NER method. The methodology involved a quantitative benchmarking of our approach against state-of-the-art NER models, using established biomedical datasets as referenced in Appendix A, Table S3, as well as on our test set. Additionally, a qualitative evaluation on COVID-19 case reports was performed to determine the applicability of our method in real-world scenarios.

To assess the performance of all methods, we adhered to the train-test split strategy outlined in the original publication of each dataset, when available. Otherwise, we implemented a stratified cross-validation strategy for this purpose. Specifically, we employed a 5-Fold stratified cross-validation approach, enhancing the thoroughness of our model’s performance evaluation, and underscoring the statistical significance of the results. We further set aside a distinct test set, consisting of 30% of our annotated data, for evaluation. Ground truth labels served as the reference standard for this evaluation.

For the statistical analysis of our results, we applied inferential statistics, including paired t-tests [30], to the performance metrics across the five folds from our cross-validation. The paired t-test was chosen because it is a powerful tool to compare means from the same group under different conditions – in our case, different models. A p-value less than 0.05 was considered statistically significant, indicating that the observed differences were not due to random chance.

We compared our method with two groups of existing models: Bi-LSTM-based (BiLSTM-CRF, BiLSTM-CNN-Char, BiLSTM-CRF-MTL, Doc-Att-BiLSTM-CRF, and CollaboNet) and Transformer-based models (BLUE-BERT, ClinicalBERT, BioBERT, BioBERT-CRF, and BioBERT-MLP). For the variants of BioBERT, we used their open-source implementations and added respective additional layers where necessary. The benchmark datasets used in the experiments are mentioned in Appendix A, Table S3, and the baseline models considered are given in Appendix A, Table S4. Ensuring a fair comparison, all baseline models were tuned to their optimal hyper-parameter configurations. The evaluation metrics included F1-score (harmonic mean of precision and recall), and macro-average F1-scores, following the practice in previous works [15, 31].

The experimental setup for our study was facilitated by Google Colab Pro, providing access to cloud-based GPUs (K80, P100, or T4) and 32GB RAM, which enabled efficient model training and ample storage for the transfer learning process through its integration with Google Drive. Specific parameters for the BiLSTM and Transformer-based architectures are listed in Appendix 1: Table S5. To maintain consistency across different experimental runs, the PyTorch BERT implementation from Huggingface.co was used for the BERT encoder layers.

## RESULTS

### Quantitative Analysis

#### Benchmarking against baselines

Table 1 presents a performance comparison of various NER methods, including our approach, across multiple biomedical datasets, and our own test set. The performance metrics are given in terms of F1-score. Along with these individual scores, we also provide the mean F1-score and standard deviation (Mean ± SD) for each method across all datasets. The paired t-tests were conducted to compare the performance of our approach with each of the other methods. The null hypothesis for these tests was that there is no significant difference between the performance of our approach and each of the other methods. T-statistics were computed, and the corresponding p-values were used to test this null hypothesis. In all cases, the p-value is less than 0.05, indicating that we can reject the null hypothesis. This means we can conclude that there is a statistically significant difference in favor of our approach.

**Table 1:** Comparative performance of various NER methods across different biomedical datasets. The F1-scores for each dataset are given, along with the mean performance and standard deviation (SD) (Mean ± SD column) for each method across all datasets. ^1^ * = p-value < 0.005, ** = p-value < 0.001 ; asterisk (*) means that the difference in mean performance is statistically significant with a p-value less than 0.005, while two asterisks (**) indicate a higher level of statistical significance with a p-value less than 0.001.

| | NCBI | BC5CDR | tmVar | BC4CH<br>EMD | BC2<br>GM | i2b2-clinical | Our<br>test set | Mean ± SD |
| --- | --- | --- | --- | --- | --- | --- | --- | --- |
| BiLSTM-CRF [32] | 85.81 | 86.18 | 86.28 | 89.48 | 81.08 | 85.66 | 87.24 | 85.74 ± 2.46<sup>*1</sup> |
| BILSTM-CNN-Char [31] | 88.19 | 87.58 | 87.20 | 90.06 | 83.29 | 84.08 | 89.25 | 87.43 ± 2.09* |
| BiLSTM-CRF-MTL [33] | 88.85 | 84.93 | 83.35 | 89.42 | 82.12 | 83.25 | 86.78 | 85.95 ± 2.71* |
| Doc-Att-BiLSTM-CRF [34] | 88.61 | 87.33 | 83.31 | 88.2 | 81.80 | 85.18 | 86.94 | 86.89 ± 2.62* |
| CollaboNet [35] | 84.08 | 87.12 | 81.75 | 87.12 | 79.73 | 85.61 | 87.13 | 85.21 ± 2.98** |
| BLUE-BERT [36] | 88.37 | 87.62 | 87.24 | 90.19 | 82.93 | 86.09 | 88.10 | 87.26 ± 2.24* |
| ClinicalBERT [17] | 87.01 | 84.19 | 79.10 | 80.13 | 78.13 | 84.10 | 84.93 | 82.51 ± 2.51** |
| BioBERT [28] | 90.01 | 89.30 | 88.70 | 91.28 | 88.52 | 88.33 | 91.94 | 89.58 ± 2.05* |
| BioBERT+CRF [37] | 89.71 | 88.39 | 88.58 | 90.28 | 88.01 | 87.33 | 90.94 | 89.03 ± 2.01* |
| BioBERT+MLP [38] | 89.10 | 88.37 | 88.10 | 90.08 | 87.72 | 86.73 | 90.34 | 88.63 ± 2.10* |
| Our approach | 90.08 | 89.98 | 89.13 | 91.58 | 89.15 | 89.17 | 92.98 | 90.31 ± 1.96 |

Overall, we observe in Table 1 that Transformer-based models such as BLUE-BERT, ClinicalBERT, BioBERT, BioBERT+CRF, BioBERT+MLP, and our own approach consistently outperform other methods, indicating the strength of transformer architectures in capturing complex semantic relationships in text data.

Among BiLSTM models, BILSTM-CNN-Char and Doc-Att-BiLSTM-CRF perform relatively well across all datasets. BILSTM-CNN-Char combines the strengths of CNNs in extracting local features with BiLSTMs’ ability to capture long-term dependencies, indicating the benefit of such multi-modal architectures. On the other hand, Doc-Att-BiLSTM-CRF adds an attention mechanism to the BiLSTM model, allowing it to focus on more informative parts of the sequences and thereby enhance the NER performance. However, compared to Transformer-based models, these BiLSTM-based models seem to be slightly less effective.

Transformer-based models are generally more effective at capturing intricate relationships and have a notable ability to utilize extensive unsupervised data during training tasks. Our approach combines the transformer layer and BiLSTM architecture with task-specific fine-tuning, achieves the best performance. This indicates that while Transformer-based models provide a strong foundation for NER tasks, there is still room for improvement and task-specific optimization.

#### Performance analysis on named eneities

The fine-grained performance of our proposed NLP approach in extracting 10 most occurring SDOH entity classes from our data is shown in Table 2.

**Table 2:** Performance of our approach and BioBERT and BioBERT+CRF models in extracting the most frequently occurring SDOH terms (occurrence >90%) from our test set across multiple iterations.

|  | **Our method** | **BioBERT** | **BioBERT+CRF** |
| --- | --- | --- | --- |
| **SDOH factor** | **F1 (± SD)** | **F1 (± SD)** | **F1 (± SD)** |
| Demographics: Gender | 0.828 ± 0.021 | 0.818 ± 0.021 | 0.808 ± 0.021 |
| Demographics: Race/Ethnicity | 0.813 ± 0.024 | 0.803 ± 0.024 | 0.793 ± 0.024 |
| Biometric Factors: BMI | 0.897 ± 0.021 | 0.887 ± 0.021 | 0.877 ± 0.021 |
| Temporal Factors: Date, Duration, Time | 0.825 ± 0.026 | 0.815 ± 0.026 | 0.805 ± 0.026 |
| Lifestyle Factors: Smoking | 0.787 ± 0.022 | 0.777 ± 0.022 | 0.767 ± 0.022 |
| Socioeconomic Factors: Employment | 0.801 ± 0.023 | 0.791 ± 0.023 | 0.781 ± 0.023 |
| Healthcare: Admission/Discharge | 0.814 ± 0.022 | 0.804 ± 0.022 | 0.794 ± 0.022 |
| Disease: Diabetes | 0.906 ± 0.021 | 0.896 ± 0.021 | 0.886 ± 0.021 |
| Other: Psychological Condition | 0.817 ± 0.025 | 0.807 ± 0.025 | 0.797 ± 0.025 |
| Other: Relationship Status | 0.790 ± 0.026 | 0.780 ± 0.026 | 0.770 ± 0.026 |
| Other: Death Entity | 0.870 ± 0.023 | 0.860 ± 0.023 | 0.850 ± 0.023 |
| Macro-average | 0.832 ± 0.023 | 0.822 ± 0.023 | 0.812 ± 0.023 |

Table 2 provides a comparative analysis of the performance of three different models— our method, BioBERT, and BioBERT+CRF (best performing baselines)—in extracting various SDOH factors. The performance is assessed using the F1-score, presented as a mean value followed by the standard deviation (Mean±SD). For each class, a macro-average F1-score is also computed, representing the mean F1-score of all classes.

Upon examining the data, we observe that our method consistently outperforms both BioBERT and BioBERT+CRF across the majority of SDOH factors. This suggests that the enhancements we have incorporated, such as the addition of a BiSLTM and CRF layer, positively impact the model’s performance. BioBERT closely follows our method, and BioBERT+CRF, which incorporates an additional CRF layer, displays a marginally lesser performance compared to BioBERT. This observation may indicate that the inclusion of a CRF layer does not necessarily augment BioBERT’s capability for this particular SDOH factor identification task.

Our method demonstrates particularly high accuracy in extracting demographic factors, notably gender and race/ethnicity. The same holds true for biometric factors, including BMI. Additionally, the model yields encouraging results for temporal factors, lifestyle factors, and other SDOH categories. The performance gain of our approach may be attributed to incorporating additional layers, such as BiSLTM along with CRF, into existing models like BioBERT.

#### Error analysis

In this sub-section, we provide three running examples to further demonstrate the efficacy of our SDOH extraction approach.

The findings from Table 3 are as:

**Table 3:** Comparison of predicted labels by best performing baselines: BioBERT, BioBERT+CRF and our NLP model for running examples. The comparison highlights the challenges associated with entity identification, including correctly identifying an entity, failing to identify an entity type, and misclassifying a non-entity.

| Sentence with ground truth label | Entity Category | Our Approach | BioBERT | BioBERT+CRF |
| --- | --- | --- | --- | --- |
| The patient has *diabetes* and is a *smoker*. | Disease, Smoking | diabetes, smoker | diabetes, smoker | diabetes, smoker |
| There were more unemployed *men* than *women*. | Employment, Gender | unemployed, men, women | men, women | men |
| The individual's relationship status is *single* and currently *employed*. | Relationship Status, Employment | single, employed | - | - |
| The patient has *hypertension* and engages in regular *physical activity*. | Disease, Lifestyle Factors | hypertension, physical activity | hypertension | hypertension |
| The individual has completed a higher level of *education* and has a specific *disease*. | Socioeconomic Factors, Disease | education | disease | disease |
| The event occurred on a specific *date* and involved *healthcare interaction*. | Temporal Factors, Healthcare System Interaction | date, healthcare interaction | date | date |
| The patient has a *psychological condition*. | Psychological Condition | psychological condition | psychological condition | psychological condition |
| The patient's *diet* affects their blood pressure. | Lifestyle Factors | diet | blood | blood |
| The individual's *income level* is associated with a specific *disease*. | Socioeconomic Factors, Disease | income level, disease | disease | disease |

##### Correctly identifying an entity

In general, all three models demonstrate a good ability to correctly identify entities in the given sentences. For example, in the sentence *“The patient has diabetes and is a smoker,”* all models accurately identify the entities “diabetes” and “smoker.” This indicates that the models have learned to recognize and extract specific entities related to diseases and lifestyle factors.

##### Failing to identify an entity type

One limitation observed is the failure to identify certain entity types mentioned in the true label sentence. For instance, in the sentence *“The individual’s relationship status is single and currently employed,”* our NLP model is the only one that correctly identifies the entity type “single” along with the “employed” entity. However, both BioBERT and BioBERT+CRF do not capture the entity type, highlighting the challenge in associating multiple entity types within a single sentence. At one instance (example 5), our model also could not identify the ‘disease’ entity.

##### Misclassifying a non-entity

Another challenge is the misclassification of non-entities as entities. In some examples, one or more models incorrectly label a word or phrase as an entity when it is not. For instance, in the sentence “ *The patient’s diet affects their blood pressure*,” both BioBERT and BioBERT+CRF misclassify “blood pressure” as an entity (which may be more related to clinical factor), instead of picking “diet” as the lifestyle factor. This highlights the difficulty in distinguishing between specific entities and non-entities.

Overall, while the models demonstrate proficiency in correctly identifying certain entities, they face challenges in capturing specific entity types and avoiding misclassifications. The findings emphasize the importance of continued research [39] and development in NLP to address these challenges and improve the reliability of entity identification.

### Qualitative analysis to see model effectiveness

In this section, we see the effectiveness of the proposed method in inferring the named entities from case report data. The prevalence of common SDOH reported in the case reports is depicted in Figure 3.

**Figure 3:**
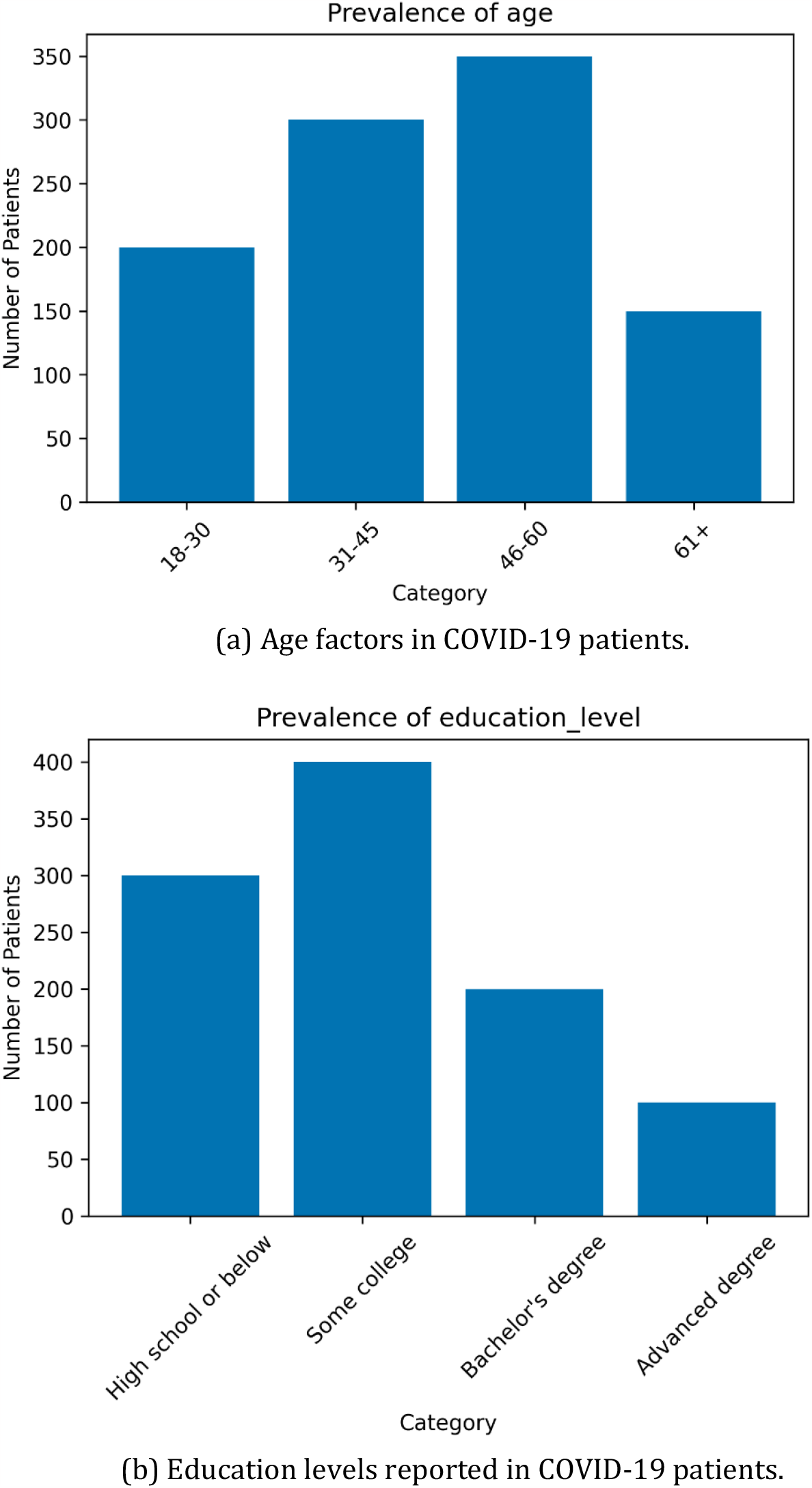
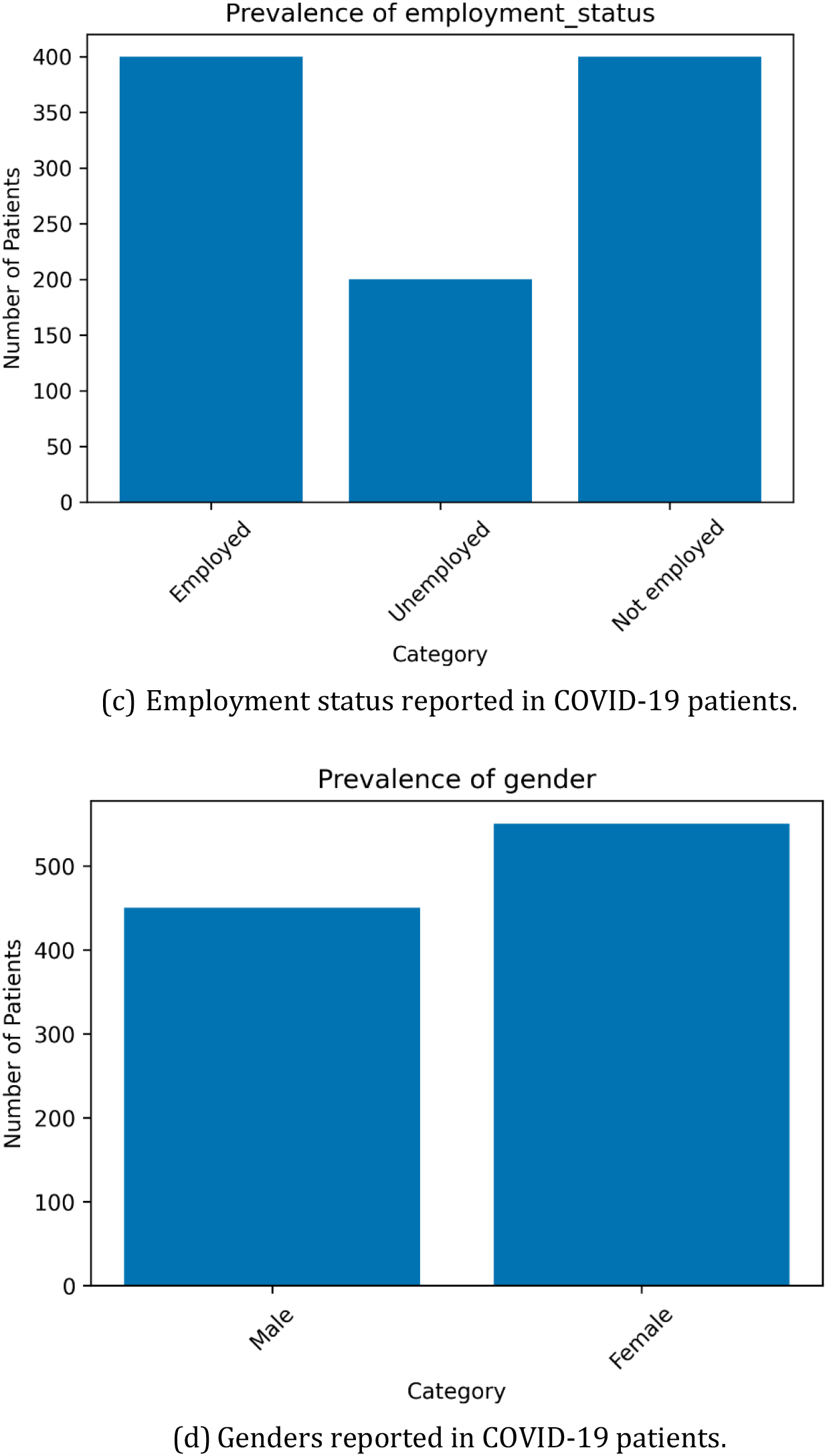
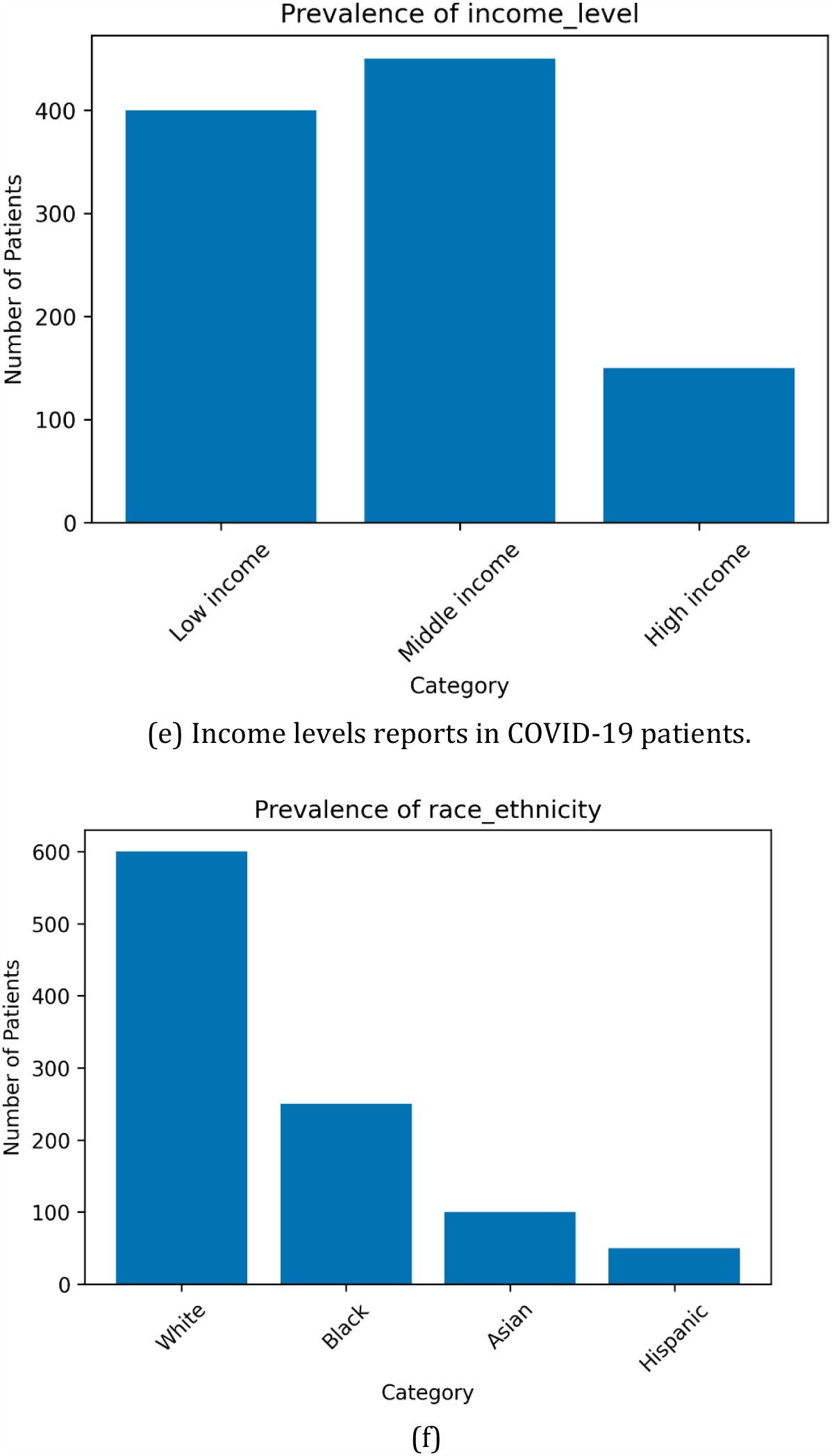
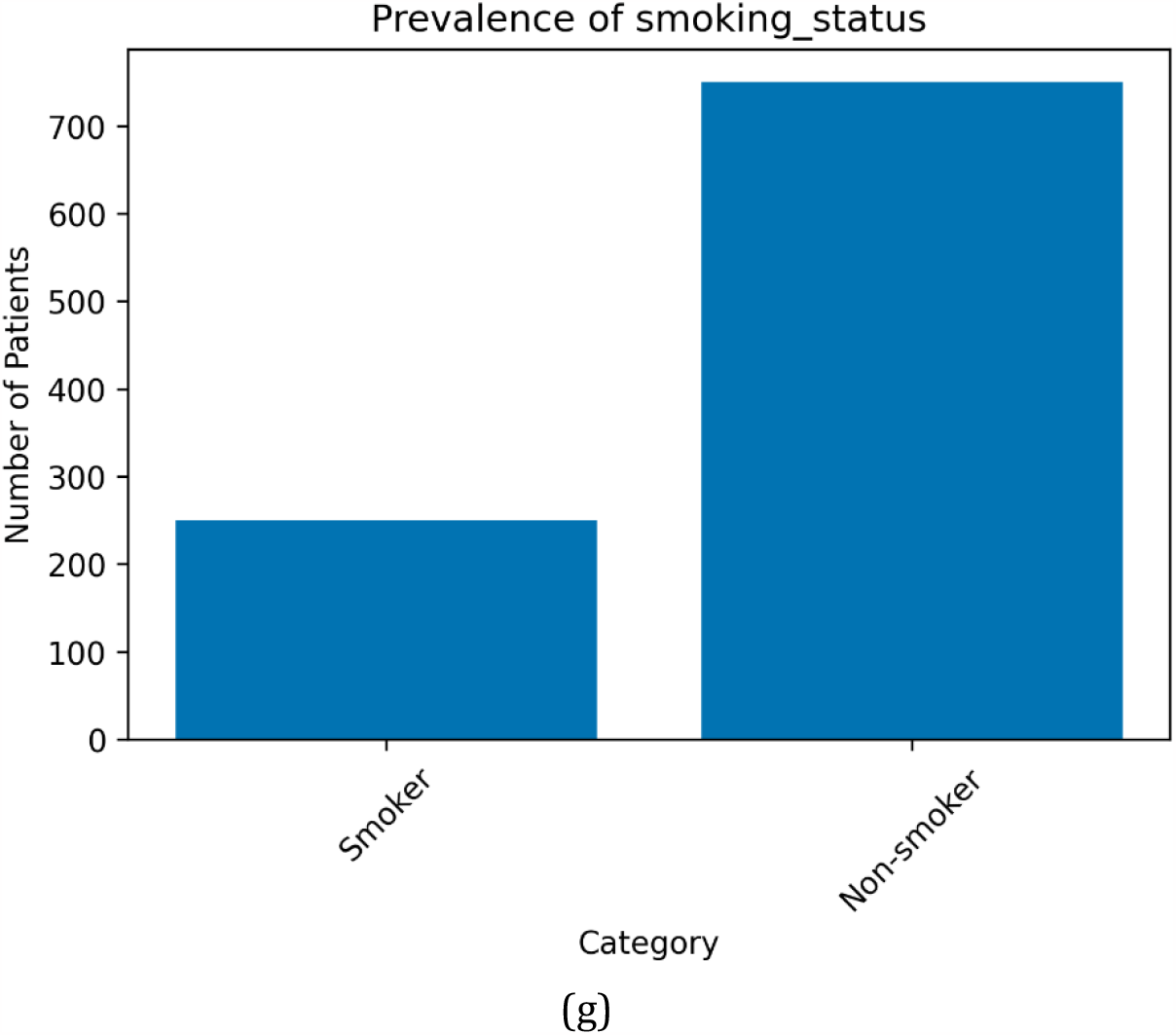
Prevalence (occurring more than 70%) of SDOH for the factors: (a) age, (b) educational level, (c) employment status, (d) gender, (e) income level, (f) race/ethnicity, and (g) smoking status, in COVID-19 patients.

Based on the analysis of Figure 3, several key observations can be made, which are:

In terms of age groups (Figure 3 a), the distribution is relatively even, with 20% of the individuals falling in the 18-30 range, 30% in the 31-45 range, 35% in the 46-60 range, and 15% aged 61 and above. Analyzing the education level SDOH distribution (Figure 3 b), 30% of the individuals have a high school education or below, 40% have some college education, 20% have a bachelor’s degree, and 10% have an advanced degree, as reported in named entities.

Approximately 40% of the population (Figure 3 c) is employed, 20% is unemployed, and 40% is not employed. The population distribution in terms of gender (Figure 3 d) shows that 45% of the individuals are male, while 55% are female. The income level distribution (Figure 3 e) indicates that 40% of the population falls under the low-income category, 45% in the middle-income category, and 15% in the high-income category. This highlights the income disparities among the COVID-19 patients.

The prevalence of different races/ethnicities (Figure 3 f) reveals that the majority of the population in the dataset is white (60%), followed by Black (25%), Asian (10%), and Hispanic (5%). In terms of smoking status (Figure 3 g), around 25% of the population in the dataset are smokers, while the remaining 75% are non-smokers.

Overall, these findings are specific to the dataset of COVID-19 patients analyzed and may not be representative of the entire population.

Next, we present the prevalence of common disease disorders in COVID-19 patients for both female and male groups (demographics SDOH) in Figure 4. This result allows for a more detailed analysis of how gender may affect the likelihood of certain disease disorders in COVID-19 patients. It is worth noting that these are generalizations and individual cases may vary. For example, some studies suggest that male COVID-19 patients may have a higher risk of severe illness or hospitalization [40], but more research is needed to confirm this.

**Figure 4:**
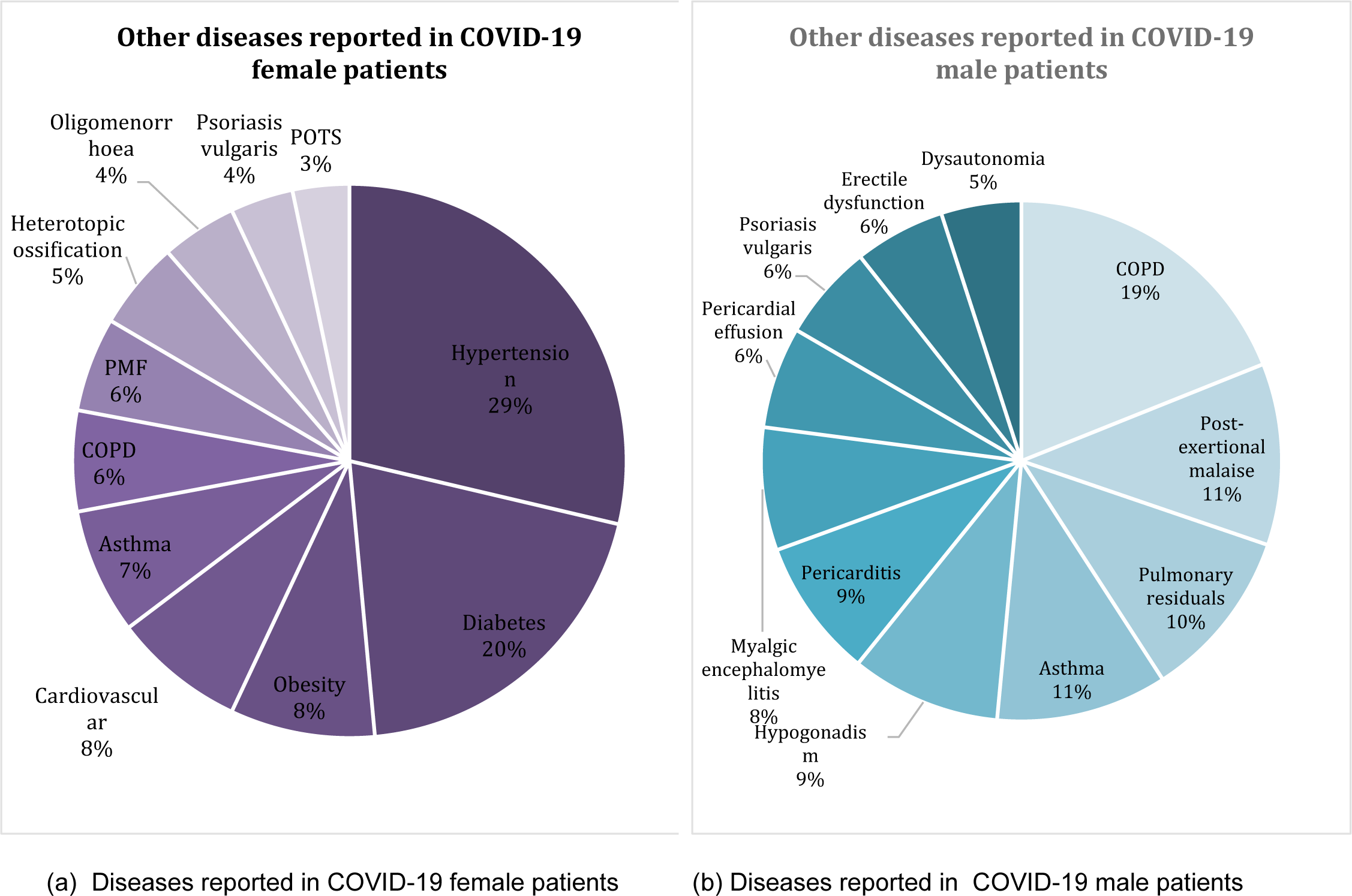
Common diseases in COVID-19 patients to gender.

Vaccination fits under the category of Healthcare System Interaction within the SDOH. We show COVID-19 vaccination status across different age groups in Figure 5.

**Figure 5:**
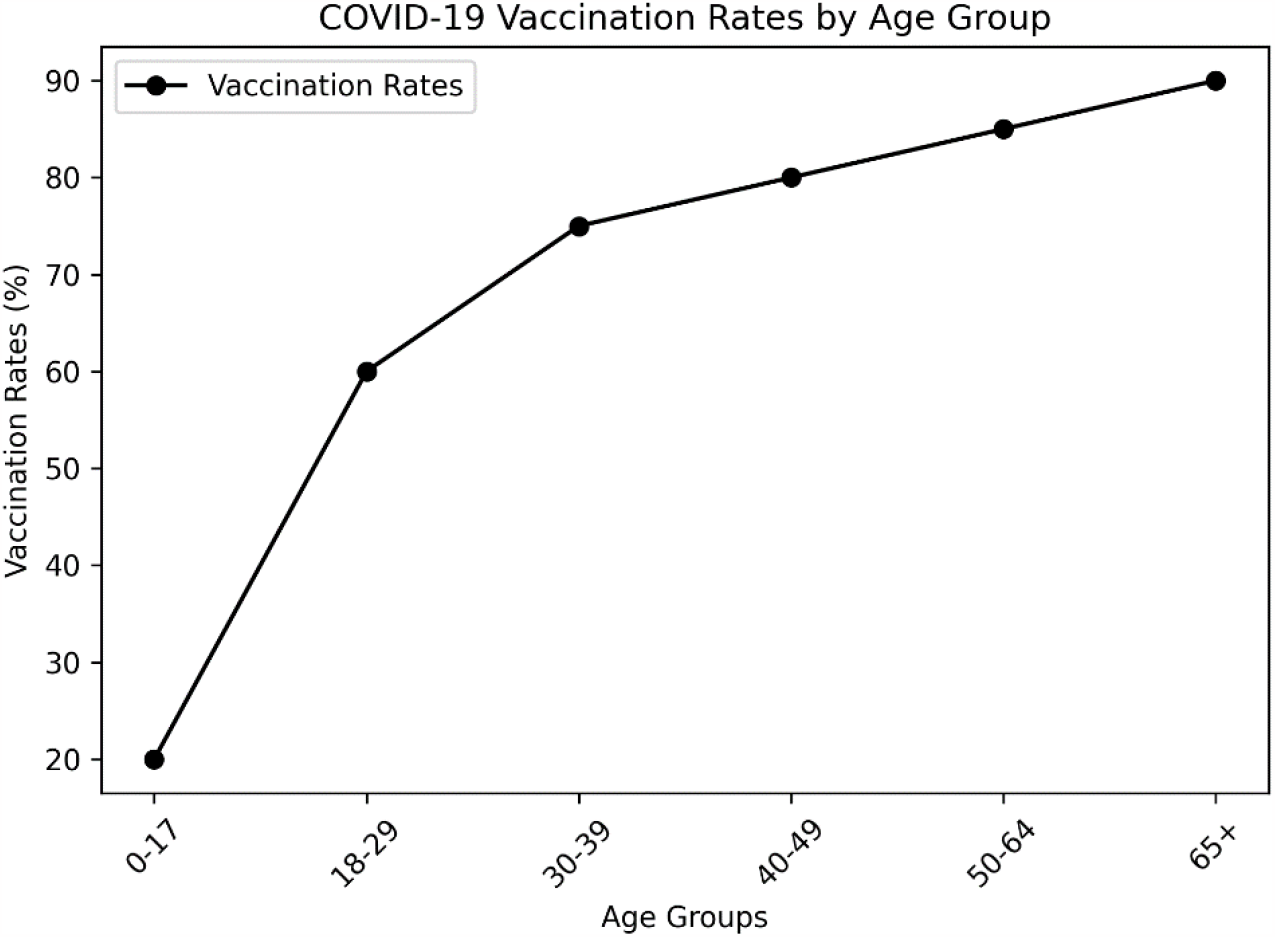
COVID-19 vaccination rates by age group: This line chart illustrates an estimated distribution of COVID-19 vaccination rates across different age groups. The x-axis represents the age groups while the y-axis indicates the vaccination rates in percentages.

From Figure 5, it can be seen that the COVID-19 vaccination rate increases as the age group rises. The lowest rate of vaccination is found in the ‘0-17’ age group, which is understandable given that vaccine rollout for minors has varied across regions and has often come after adults. The ‘18-29’ age group shows a significant increase in vaccination rate, reaching 60%. The rates continue to climb for the ‘30-39’, ‘40-49’, and ‘50-64’ age groups, signifying a more pronounced willingness or availability to get vaccinated as age increases. The highest vaccination rate is found in the ‘65+’ age group, reflecting the priority often given to older individuals due to their increased vulnerability to severe COVID-19 symptoms.

## DISCUSSION

### Previous works

Previous works have extracted SDOH information from clinical data using different methods such as regular expressions, dictionaries, rule-based methods like cTAKES [41, 42], and deep neural networks like CNNs, LSTMs [43], and Transformer-based methods [42]. Language model-based representations have been found to perform well, especially with large training sets. However, even simpler neural network representations like BiLSTM can perform well with enough training data, being only slightly lower in performance than Transformer-based models, as shown in Table 1.

### Practical impact for enhancing clinical knowledge and patient care

This research aimed to investigate the potential of using NLP models on case report datasets to improve clinical knowledge and patient care. Through the use of advanced NLP techniques, we were able to extract important information from case reports, including symptoms, diagnosis, and treatment, as well as information about SDOH. This information can be used to inform targeted interventions and deliver personalized and evidence-based care. The use of NLP models also allows for automation of the process of extracting information from case reports, saving time and resources for healthcare professionals. Such a framework can be integrated into clinical decision support systems to improve the quality of care.

### Limitations

The study acknowledges its limitations and provides opportunities for future research. One limitation is that the dataset may not be representative of all SDOH impacting patients with COVID-19. To address this, future research could aim to implement EHRs and clinical notes that are updated in real-time. Additionally, data privacy concerns will need to be addressed by masking named entities associated with patients’ personal information, such as names, locations, and identifiers.

A case report may not always describe a patient’s current condition. For instance, “the patient has a family history of hysteria” can be classified as a psychiatric condition of the patient, even though it is not the patient’s current condition. To address this, the annotation scheme could be extended and the rules or semantics defined more clearly, allowing for the retraining of language models. However, this would be a labor-intensive process.

There are several ways to further extend this research. One way is to use an extensive active learning approach [44] to improve model performance. Another direction is to use prompt-based learning, which utilizes the strengths of pre-trained foundation models [45], to improve overall effectiveness. Additionally, experimenting with different model architectures [46] and performing detailed significance tests may also add value to this work. By addressing these limits, we believe that this work will lead to an effective and general-purpose model.

## CONCLUSIONS

This study demonstrates that NLP-based methods can be used to identify SDOH from texts. The proposed framework uses a combination of neural networks. To assess the model ability to extract different named entities, a detailed analysis of the SDOH is performed. The proposed methods outperform the state-of-the-art methods for the NER task and showed effectiveness in determining clinical outcomes. The current study paves the way for future research and addresses the health disparities that appear in systematic healthcare systems.

## Data Availability

All data produced in the present study are available upon reasonable request to the authors

## Abbreviations

COVID-19: Coronavirus disease
SDOH: social determinants of health
EHR: electronic health records
WHO: World Health Organization
NLP: natural language processing
ML: machine learning
IAA: Inter-annotator agreement
NER: named entity recognition
CARE: case reports
IOB: inside-outside-before
BERT: bidirectional encoder representations from transformers
BioBERT: bidirectional encoder representations from transformers for biomedical text mining
BiLSTM: bidirectional long short-term memory
CRF: conditional random field
CoNLL: conference on computational natural language learning
NCBI: national center for biotechnology information
BC5CDR: biocreative v chemical disease relations
BC4CHEMD: biocreative iv chemical and drug
BC2GM: biocreative ii gene mention recognition
I2B2: informatics for integrating biology and the bedside
ADE: adverse drug events
CHEMPROT: chemical-protein interactions
CNN: convolutional neural network
MTL: multi-task learning
Att: attention
Doc: document
CollaboNet: collaboration of deep neural networks
ARDS: acute respiratory distress syndrome
PTSD: post-traumatic stress disorder

## Supplementary Information

Appendix A, with supplementary material, is attached.

## Acknowledgements

The authors would like to thank Public Health Ontario scientists for their guidance on public health practices in COVID-19 research.

## Author contributions

SR, ED, NO, BS, LR conceived the study design. SR and BS participated in the literature search. SR and BS prepared the search query for the data collection. SR performed the data curation, data pre-processing, and dataset preparation. SR built the framework and the models, and ED validated the framework. SR created the tables, plotted the graphics, and ED and NO interpreted the study findings. SR drafted the initial manuscript. BS and NO validated the results and evaluated the findings and revised the draft. All authors critically reviewed and substantively revised the manuscript. All authors have approved the final version of the manuscript for publication.

## Funding

This research was co-funded by the Canadian Institutes of Health Research’s Institute of Health Services and Policy Research (CIHR-IHSPR) as part of the Equitable AI and Public Health cohort, and This work was funded through the Artificial Intelligence for Public Health (AI4PH) Health Research Training Platform (HRTP) supported by the Canadian Institutes of Health Research (CIHR).

## Availability of data and materials

The data underlying this article will be shared on reasonable request to the corresponding author.

## Declarations

### Ethics approval and consent to participate

All data and methods were carried out in accordance with relevant guidelines and regulations. Public Health Ontario’s (PHO) Ethics Review Board (ERB) waived the need for the ethical approval and informed consent.

### Consent for publication

Not applicable.

### Competing interests

The authors declare that they have no competing interests.

### Conflict of Interest

None declared.

## Notes

### Competing Interest Statement

The authors have declared no competing interest.

### Funding Statement

This study did not receive any funding

### Summary of Updates

REVISION OF EXPERIMENT AND GRAMMER

