## SUPPLEMENTARY for "Discovering Social Determinants of Health from Case Reports using Natural Language Processing: Algorithmic Development and Validation"

^1^ Public Health Ontario (PHO), Toronto, ON, Canada.

^2^ Dalla Lana School of Public Health, University of Toronto, Toronto, ON, Canada.

^3^ Institute of Health Policy, Management and Evaluation (IHPME), University of Toronto, Toronto, ON, Canada.

Corresponding Author:

Shaina Raza, PhD

**Appendix A**

**Table S1**: *Search Query*

("2022/01/01"[Date - Publication] : "2022/06/30"[Date - Publication]) AND

English[Language] AND

(("6"[Age] : "12"[Age]) OR

("13"[Age] : "18"[Age]) OR

("19"[Age] : "44"[Age]) OR

("45"[Age] : "64"[Age]) OR

("65"[Age] : "120"[Age])) AND

"Case Reports"[Publication Type]

**Table S2:** *Entities*

**To better organize these entities, we can categorize them into distinct clinical and non-clinical (including SDOHs) categories.**

**Non-Clinical Entities (including SDOHs)**

1. *Demographics*: Gender, Age, Race/Ethnicity
2. *Biometric factors*: Height, Weight
3. *Temporal Factors*: Date, Relative Date, Duration, Time
4. *Lifestyle Factors*: Smoking, Alcohol, Substance Use
5. *Socioeconomic Factors*: Employment
6. *Healthcare System Interaction:* Admission/Discharge, Vaccination
7.

**Clinical Entities**

1. Cardiovascular Conditions: Heart Disease, Hypertension, Hyperlipidemia
2. Metabolic Conditions: Diabetes, Obesity, BMI
3. Renal Conditions: Kidney Disease
4. Oncological Conditions: Oncological
5. Respiratory Conditions: Respiration
6. Injury or Poisoning
7. Psychological Conditions
8. Symptoms
9. Internal Body Parts
10. External Body Parts
11. Treatment Procedures: Treatment, Procedure
12. Clinical Measures: Blood Pressure, Dosage, Pulse, Test
13. Medications: Drug Name, Vaccine
14. Clinical Department
15. Death Entity

**Figure S1:** Named entities extracted from the case report (1).

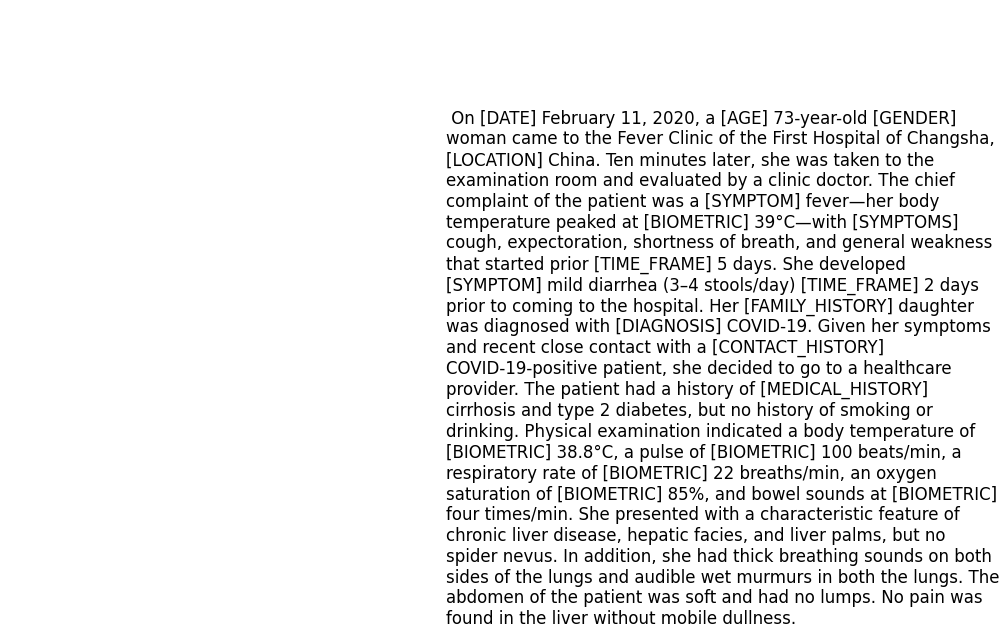

**Table S3***: Benchmark datasets used.*

| **Corpus** | **Entity types** | **Data size** |
| --- | --- | --- |
| NCBI-Disease ^47^ | Diseases | 793 Pubmed abstracts |
| BC5CDR ^48^ | Diseases and diseases | 1500 Pubmed articles |
| tmVar ^50^ | Disease-related mutation | 500 PubMed articles |
| BC4CHEMD ^51^ | Chemicals | 10,000 Pubmed abstracts |
| BC2GM ^52^ | Gene/Proteins | 20,000 sentences |
| JNLBPA ^53^ | Genes, proteins | 2404 abstracts |
| i2b2-Clinical ^54^ | Problem, Treatment, and Test. | 426 discharge summaries |

**Table S4***: Baseline methods used.*

| **Baseline method** | **Description** |
| --- | --- |
| BiLSTM-CRF (2) | This is a neural network model that combines Bidirectional Long Short-Term Memory (BiLSTM) and Conditional Random Fields (CRF) for sequence prediction tasks like NER. |
| BILSTM-CNN-Char (3) | This model combines BiLSTM, Convolutional Neural Network (CNN), and Character Embedding for more effective sequence prediction tasks. It leverages both word-level and character-level representations. |
| BiLSTM-CRF-MTL (4) | This is a model that combines BiLSTM, CRF, and Multi-Task Learning (MTL). MTL helps the model to learn and generalize better by training it on multiple related tasks concurrently. |
| Doc-Att-BiLSTM-CRF (5), | This model uses Document Attention mechanism along with BiLSTM and CRF for sequence prediction tasks. The attention mechanism helps the model to focus on more relevant parts of the input data. |
| CollaboNet (6) | This model uses Document Attention mechanism along with BiLSTM and CRF for sequence prediction tasks. The attention mechanism helps the model to focus on more relevant parts of the input data. |
| BLUE-BERT (7) | This is a version of the BERT model pre-trained on both clinical and biomedical text corpus for improved performance on healthcare related Natural Language Processing tasks. |
| ClinicalBERT (8) | This is a variant of the BERT model pre-trained specifically on clinical text to perform various tasks in medical natural language processing. |
| BioBERT (9) | This is a domain specific language representation model pre-trained on large-scale biomedical corpora. It is designed for biomedical text mining tasks such as disease name recognition, species recognition, and chemical compound and drug name recognition. |
| BioBERT+CRF (10) | This model leverages the powerful language representation abilities of BioBERT and combines it with the sequence prediction capabilities of CRF for tasks like ner in biomedical text. |
| BioBERT+MLP (11) | This model uses BioBERT for extracting robust language representations and a Multi-Layer Perceptron (MLP) for final prediction tasks. The combination helps to adapt to various tasks in biomedical NLP. |

**Table S5:** *General Hyperparameters used along with best value and range in parenthesis.*

| **Hyperparameter** | **Value** |
| --- | --- |
| LSTM state size | 200 [200 - 300] |
| dropout rate | 0.5 [0.2 - 0.7] |
| Epochs | 40 [20- 80] |
| Batch size | 16 [8 - 128] |
| Learning rate (lr) | 1.e-05 [1.e-9 – 1.e-2] |
| lr decay coefficient (po) | 0.005 [0.001, 0.01] |
| Warmup steps | 10,000 |
| Optimizer | ADAM (12), β1=0.9 and β2=0.999 |
| Word dimension | 300 [50 – 450] |
| Hidden size LSTM | 300 |
| Gradient clipping | 5.0 |
| The fine-tuning transformer-based architectures (BioBERT and others) are maximum sequence length of 128, number of layers as 12, number of attention heads also 12 and embedding size as 768. For different datasets, the fine-tuning takes different hours (2 hours, 3 hours, 4 hours and 10 hours for our dataset). In the NER task, we fixed the length of sentences to 512. Other hyperparameters used in the paper are given in Table 2. | |

1.	Zhou J, Jiang D, Wang W, Huang K, Zheng F, Xie Y, et al. Case Report: Clinical Features of a COVID-19 Patient With Cirrhosis. Front Med. 2021;8.

2.	Lample G, Ballesteros M, Subramanian S, Kawakami K, Dyer C. Neural architectures for named entity recognition. arXiv Prepr arXiv160301360. 2016;

3.	Chiu JPC, Nichols E. Named Entity Recognition with Bidirectional LSTM-CNNs. Trans Assoc Comput Linguist. 2016;4(2003):357–70.

4.	Wang X, Zhang Y, Ren X, Zhang Y, Zitnik M, Shang J, et al. Cross-type biomedical named entity recognition with deep multi-task learning. Bioinformatics. 2019;35(10):1745–52.

5.	Xu K, Yang Z, Kang P, Wang Q, Liu W. Document-level attention-based BiLSTM-CRF incorporating disease dictionary for disease named entity recognition. Comput Biol Med. 2019;108:122–32.

6.	Yoon W, So CH, Lee J, Kang J. Collabonet: collaboration of deep neural networks for biomedical named entity recognition. BMC Bioinformatics. 2019;20(10):55–65.

7.	Peng Y, Yan S, Lu Z. Transfer learning in biomedical natural language processing: an evaluation of BERT and ELMo on ten benchmarking datasets. arXiv Prepr arXiv190605474. 2019;

8.	Huang K, Altosaar J, Ranganath R. ClinicalBERT: Modeling Clinical Notes and Predicting Hospital Readmission. arXiv Prepr arXiv190405342 [Internet]. 2019; Available from: http://arxiv.org/abs/1904.05342

9.	Lee J, Yoon W, Kim S, Kim D, Kim S, So CH, et al. BioBERT: A pre-trained biomedical language representation model for biomedical text mining. Bioinformatics [Internet]. 2020;36(4):1234–40. Available from: https://github.com/dmis-lab/

10.	Sun C, Yang Z, Wang L, Zhang Y, Lin H, Wang J. Biomedical named entity recognition using BERT in the machine reading comprehension framework. J Biomed Inform [Internet]. 2021;118(May):103799. Available from: https://doi.org/10.1016/j.jbi.2021.103799

11.	Luo X, Gandhi P, Storey S, Huang K. A deep language model for symptom extraction from clinical text and its application to extract covid-19 symptoms from social media. IEEE J Biomed Heal Informatics. 2021;26(4):1737–48.

12.	Kingma DP, Ba JL. Adam: A method for stochastic optimization. In: 3rd International Conference on Learning Representations, ICLR 2015 - Conference Track Proceedings. International Conference on Learning Representations, ICLR; 2015.
